# Variation in SARS-CoV-2 free-living survival and environmental transmission can modulate the intensity of emerging outbreaks

**DOI:** 10.1101/2020.05.04.20090092

**Authors:** C. Brandon Ogbunugafor, Miles D. Miller-Dickson, Victor A. Meszaros, Lourdes M. Gomez, Anarina L. Murillo, Samuel V. Scarpino

## Abstract

Variation in free-living, microparasite survival can have a meaningful impact on the ecological dynamics of established and emerging infectious diseases. Nevertheless, resolving the importance of environmental transmission in the ecology of epidemics remains a persistent challenge, requires accurate measuring the free-living survival of pathogens across reservoirs of various kinds, and quantifying the extent to which interaction between hosts and reservoirs generates new infections. These questions are especially salient for emerging pathogens, where sparse and noisy data can obfuscate the relative contribution of different infection routes. In this study, we develop a mechanistic, mathematical model that permits both direct (host-to-host) and indirect (environmental) transmission and then fit this model to empirical data from 17 countries affected by an emerging virus (SARS-CoV-2). From an ecological perspective, our model highlights the potential for environmental transmission to drive complex, non-linear dynamics during infectious disease outbreaks. Summarizing, we propose that fitting such models with environmental transmission to real outbreak data from SARS-CoV-2 transmission highlights that variation in environmental transmission is an underappreciated aspect of the ecology of infectious disease, and an incomplete understanding of its role has consequences for public health interventions.

## Introduction

The ecology of infectious disease has provided a theoretical basis for understanding how interactions between microbes, their hosts, and environments, shape the severity of epidemics. Mathematical modeling methods offer means of testing a systems perspective such that we can explore—if even theoretically—the consequences of underappreciated routes of transmission in epidemics. This is useful in the case of emerging diseases, where it may be challenging to fully resolve all of the features of an epidemic, including the influence of different routes of transmission on key features of an epidemic. That is, even though there is a large literature on environmental transmission (or “fomite” transmission), relatively few studies have explored how particulars aspects of environmental transmission—the nature, composition, and frequency of environmental reservoirs—can influence disease dynamics.

Lack of clarity regarding of the signature of variation in environmental transmission is especially notable in emerging diseases, where data sets are elusive, sparse, or noisy. For example, Severe Acute Respiratory Syndrome Coronavirus 2 (SARS-CoV-2), the etiological agent of coronavirus disease 2019 (COVID-19), has caused one of the most devastating pandemics of the last century. The complex set of epidemiological characteristics defining COVID-19 outbreaks presents a number of challenges for controlling this disease. As a consequence, countries have achieved varying levels of success in reducing transmission and protecting vulnerable populations, often with dramatic variation from setting to setting in the epidemic growth rate, intensity, and/or severity. The basic reproductive number (ℛ_*0*_) (1-3), fatality rate (4-5), incubation period (4, 6-8), transmission interval (9), prevalence of super-spreading events (10-11) and other relevant aspects of COVID-19 epidemiology provide a mechanistic window into how SARS-CoV-2 is transmitted in different settings. However, one feature of SARS-CoV-2 transmission that was validated in laboratory settings, but whose epidemiological role remains highly controversial, is SARS-CoV-2 free-living survival (12-14).

Specifically, while several laboratory and epidemiological findings have suggested that environmental transmission may play a role in some settings (11, 13-16), none have fully investigated how this route of transmission may influence features of outbreaks, and have cast doubt about its relevance as a significant mode of transmission at all (17). These unknowns notwithstanding, the availability of SARS-CoV-2 data make it a valuable model for examining the particulars of environmental transmission in emerging outbreaks.

In this study, we use confirmed case data and laboratory and epidemiologically validated parameters to develop a mechanistic transmission model. Using this, we evaluate the potential for variability in environmentally-mediated transmission to explain variability in various features associated with the intensity of outbreaks. This framework includes parameters corresponding to the transmission of the virus from both presymptomatic/asymptomatic and clinical (symptomatic) carriers of virus, and the possibility that susceptible hosts can acquire infection through environmental reservoirs. We examine how outbreak dynamics can be influenced by differences in viral free-living survival that include empirical values for survival on various abiotic reservoirs (e.g. aerosols, plastic, copper, steel, cardboard) (12).

Specifically, our findings highlight the need for an empirically-informed, mechanistic understanding of the ecology of emerging viral outbreaks—including the particulars of how free-living virus survives on different abiotic surfaces, and in aerosol form—should be a greater focus in emerging infectious disease outbreaks, as lack of clarity regarding the specifics of this route can obfuscate important details of outbreak dynamics.

## Materials and Methods

### A *Waterborne, Abiotic*, and other *Indirectly Transmitted* (WAIT) model for the dynamics of emergent viral outbreaks

Several models have been engineered to explore aspects of COVID-19 dynamics. For example, models have been used to investigate the role of social distancing (2, 18), social mixing (19), the importance of undocumented infections (20), the role of mobility in the early spread of disease in China (21), and the potential for contact tracing as a solution (22). Only a few notable models of SARS-CoV-2 transmission incorporate features of indirect or environmental transmission (15, 16, 22), and none consider the dynamical properties of viral free-living survival in the environment. Such a model structure would provide an avenue towards exploring how variation in free-living survival influences disease outbreaks. Environmental transmission models are aplenty in the literature and serve as a theoretical foundation for exploring similar concepts in newer, emerging viruses (23-32).

Here, we parameterize and validate an SEIR-W model: Susceptible (*S*), Exposed (*E*), Infectious (*I*), Recovered (*R*), and WAIT (*W*) model. Here *W* represents the environmental component of the transmission cycle during the early stage of the SARS CoV-2 pandemic. This model is derived from a framework previously developed called the “WAIT” modeling framework—which stands for *Waterborne, Abiotic*, and other *Indirectly Transmitted—*that incorporates an environmental reservoir where a pathogen can sit and *wait* for hosts to interact with it (33, 34).

### Building the SEIR-W model framework for SARS-CoV-2

Here *W* represents the environmental component of the early stage of the SARS CoV-2 pandemic (Fig. S1). This environmental compartment refers to reservoirs that people may have contact with on a daily basis, such as doorknobs, tables, chairs, mail packages, and non-circulating air indoors. The *W* compartment of our model represents the fraction of these environmental reservoirs that house some sufficiently transmissible amount of infectious virus. We emphasize that the *W* compartment is meant to only represent reservoirs that are common sites for interaction with people. Thus, inclusion of the *W* compartment allows us to investigate the degree to which the environment is infectious at any given point, and its impact on the transmission dynamics of SARS CoV-2.

Model parameters are described in detail in Table 1. The system of equations in the proposed mathematical model corresponding to these dynamics are defined in equations 1-6:

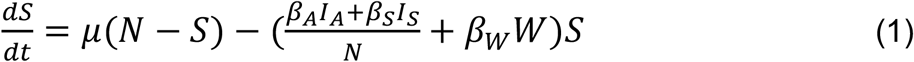

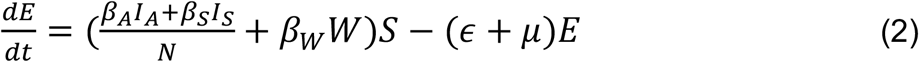

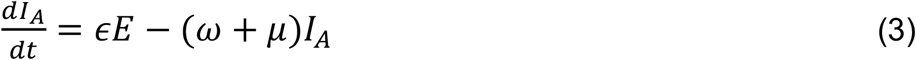

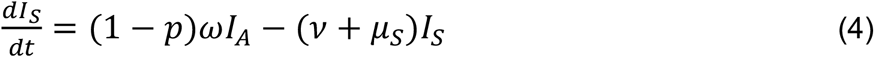

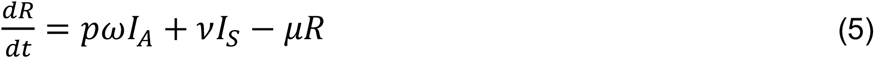

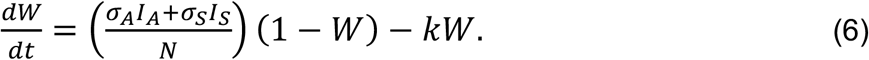

**Table 1.** Model population definitions and initial values denoted with subscript 0 for each state variable. Here we present definitions for the population groups represented by each compartment as well as their initial values. The initial value of the *S* and *I*_*S*_ populations vary by country, as shown in Table 2. We take the initial value of the *I*_*A*_ population to be the same as the initial value of symptomatic individuals as a conservative estimate. The initial value of the *E* population is computed by assuming that all initially-infected people (*I*_*A0*_ *+ I*_*S0*_) have exposed the virus to approximately ℛ_*0*_ (≈ 2.5) other people.

| Symbols | Initial Values | Units | Definitions | Sources |
| --- | --- | --- | --- | --- |
| $S_0$ | Varies by country | people | Susceptible individuals | Vary |
| $E_0$ | $\mathcal{R}_0 \cdot (I_{A0} + I_{S0})$ | people | Exposed individuals | Deduced |
| $I_{A0}$ | $I_{S0}$ | people | Asymptomatic individuals | Deduced |
| $I_{S0}$ | Varies | people | Symptomatic individuals | Deduced |
| $Rec_0$ | 0 | people | Recovered individuals | Deduced |
| $W_0$ | 1% | unitless | % of viruses in environment | Deduced |

### Infection trajectories

In addition to including a compartment for the environment (W), our model also deviates from traditional SEIR form by splitting the infectious compartment into an *I*_*A*_*-*compartment (*A* for asymptomatic), and an *I*_*S*_*-*compartment (*S* for symptomatic). As we discuss below, including asymptomatic (or sub-clinical) transmission is both essential for understanding how we environmental—as opposed to simply unobserved or hidden—transmission effects the ecological dynamics of pathogens and also for analyzing SARS-CoV-2. The former represents an initial infectious stage (following the non-infectious, exposed stage), from which individuals will either move on to recovery directly (representing those individuals who experienced mild to no symptoms) or move on to the *I*_*S*_*-*compartment (representing those with a more severe response). Finally, individuals in the *I*_*S*_*-*compartment will either move on to recovery or death due to the infection. This splitting of the traditional infectious compartment is motivated by mounting evidence of asymptomatic transmission of SARS CoV-2 (20, 36-39). Thus, we consider two trajectories for the course of the disease, similar to those employed by (18): (1) *E* → *I*_*A*_ → *R* and (2) *E* → *I*_*A*_ → *I*_*S*_ → *R (or death)*. More precisely, once in the *E* state, an individual will transition to the infectious state *I*_*A*_, at a per-person rate of ε. A proportion *p* will move from *I*_*A*_ to the recovered state *R* (at a rate of *p* ω). A proportion *(1 - p)* of individuals in the *I*_*A*_ state will develop more severe systems and transition to *I*_*s*_ (at a rate of *(1 - p)* ω). Individuals in the *I*_*s*_ state recover at a per-person rate of *v* or die at a per-person rate *μ*_*S*_. In each state, normal mortality of the individual occurs at the per-person rate *μ* and newly susceptible (*S*) individuals enter the population at a rate *μN*. The important differences between these two trajectories are in how likely an individual is to move down one path or another, how infectious individuals are (both for people and for the environment), how long individuals spend in each trajectory, and how likely death is along each trajectory.

### Clarification on the interactions between hosts and reservoirs

The model couples the environment and people in two ways: (1) people can deposit the infectious virus to environmental reservoirs (e.g. physical surfaces, and in the case of aerosols, the ambient air) and (2) people can become infected by interacting with these reservoirs (infecting the people). While the gist of our study is focused on physical surfaces, we also include data and analysis of SARS-CoV-2 survival in aerosols. While aerosols likely play a significant role in person-to-person transmission, they also facilitate an indirect means of transmitting. For example, in indoor settings an infected individual can deposit infectious SARS-CoV-2 into aerosols, which can then infect other individuals in an indoor setting (41, 42). These can then remain suspended in the air, whereby other individuals can become infected without ever having to be in especially close physical proximity to the aerosol emitter (only requires that they interact with the same stagnant air, containing infectious aerosol participates). In this setting, the free-living survival of SARS-CoV-2 in aerosol form is a very relevant parameter in determining disease dynamics.

Environmental reservoirs infect people through the *β*_*W*_ term (equations 1 and 2), a proxy for a standard transmission coefficient, corresponding specifically to the probability of successful infectious transmission from the environment reservoir to a susceptible individual (the full rate term being *β*_*W*_*W·S*). Hence, the *β*_*W*_ factor is defined as the fraction of people who interact with the environment daily, per fraction of the environment, times the probability of transmitting infection from environmental reservoir to people. The factor *β*_*W*_*W* (where *W* is the fraction of environmental reservoirs infected) represents the daily fraction of people that will interact with the infected portion of the environment and become infected themselves. The full term *β*_*W*_*W·S* is thus the total number of infections caused by the environment per day.

In an analogous manner, we model the spread of infection *to* the environment with the two terms *σ*_*A*_ *I*_*A*_*·(1 - W) / N* and *σ*_*S*_ *I*_*S*_*·(1 - W) / N* representing deposition of infection to the environment by asymptomatic individuals, in the former, and symptomatic individuals, in the latter. In this case, *σ*_*A*_ (and analogously for *σ*_S_) gives the fraction of surfaces/reservoirs that interact with people at least once per day, times the probability that a person (depending on whether they are in the *I*_*A*_ or the *I*_*S*_ compartment) will deposit an infectious viral load to the reservoir. Thus, *σ*_*A*_ *I*_*A*_ */ N* and *σ*_*S*_ *I*_*S*_ */ N* (where *N* is the total population of people) represent the daily fraction of the environment that interacts with asymptomatic and symptomatic individuals, respectively. Lastly, the additional factor of *(1 - W)* gives the fraction of reservoirs in the environment that have the potential for becoming infected, and so *σ*_*A*_ *I*_*A*_*·(1 - W) / N* (and analogously for *I*_*S*_) gives the fraction of the environment that becomes infected by people each day. We use *W* to represent a fraction of the environment, although one could also have multiplied the *W* equation by a value representing the total number of reservoirs in the environment (expected to remain constant throughout the course of the epidemic, assuming no intervention strategies).

### Parameter values estimation

Table 1 displays information on the population definitions and initial values in the model. Tables 2 and 3 contain the fixed and estimated values and their sources (respectively). Because this model iteration is relatively underexplored with regards to COVID-19, we have worked to justify its use in various ways. The model’s estimated parameters are based on model fits to 17 countries with the highest cumulative COVID-19 cases (of the 181 total countries affected) as of 03/30/2020, who have endured outbreaks that had developed for at least 30 days following the first day with ≥10 cumulative infected cases within each country (40) (See supplementary information Tables S1 – S3). In addition, we compare country fits of the SEIR-W model to fits with a standard SEIR model. Lastly, we compare how various iterations of these mathematical models compare to one another with regards to the general model dynamics. For additional details, see the supplementary information.

**Table 2.** Fixed parameter values estimated based on available published literature. These estimated values derived from the existing COVID-19 and SARS-CoV-2 literature.

| Symbols | Values | Units | Definitions | Sources |
| --- | --- | --- | --- | --- |
| $\mu$ | 80.3 x 365 | 1/day | Natural Death Rate (Reciprocal of the average life expectancy of 17 countries sampled) | (40, 47) |
| $\mu_s$ | 0.00159 | 1/day | Infected death rate | (4, 35) |
| $\eta$ | 5.5 | days | Incubation Period | (43) |
| $1/\omega$ | $\eta - \varepsilon^{-1}$ | days | Expected time in the asymptomatic state | Fitted and dependent on $\eta$ |
| $\nu$ | 0.031 | 1/day | Recovery rate (Average of 3 to 6 weeks) | (43) |
| $p$ | 0.956 | unitless | Fraction that move along the “mild” recovery track | (2) |
| $k$ | 0.649 | 1/day | Viral decay rate in environment (using average of all material values, wood, steel, cardboard, plastic) | (12) |

**Table 3.** Estimated parameter values, averaged across countries. Here we provide a table of the average values of the fitted parameters used in this model. These averages are taken across all of the selected 17 countries. See supplementary information for more details on country data and parameter estimation.

| Symbols | Average values (SEIR-W) | Standard Deviation (SEIR-W) | Average values (SEIR) | Standard Deviation (SEIR) | Units | Definitions |
| --- | --- | --- | --- | --- | --- | --- |
| $\beta_A$ | 0.550 | 0.345 | 0.429 | 0.751 | 1/day | (Contact rate of people with people) x (transmission probability of people to people by an asymptomatic person) |
| $\beta_S$ | 0.491 | 1.260 | 8.019 | 5.972 | 1/day | (Contact rate of people with people) x (transmission probability of people to people by I-person) |
| $\beta_W$ | 0.031 | 0.039 | 0.0 | -- | 1/day | (Contact rate of person with environment) x (transmission probability of environment to people) |
| $\sigma_A$ | 3.404 | 6.662 | 0.0 | -- | 1/day | (Contact rate of person with environment) x (probability of shedding by asymp.-person to environment) |
| $\sigma_S$ | 13.492 | 18.849 | 0.0 | -- | 1/day | (Contact rate of person with environment) x (probability of shedding by symp.-person to environment) |
| $1/\epsilon$ | 2.478 | 1.325 | 2.381 | 2.249 | days | Average number of days before infectious |

### Estimation of fixed parameters

There are 6 fixed parameters, 6 fitted parameters, and one parameter (ω) dependent on the values of one of the fixed parameters (*η*) and one of the fitted parameters (ε). These fixed parameters are *η, μ, μ*_S_, *v, k*, & *p*. The first, *η*, is the incubation period) (6, 43), and we assume that the expected time in the *E* state (1/ε) and the expected time in the *I*_*A*_ state (1/ω) sums to *η*, i.e. *η* = 1/ε + 1/ω. Fixing *η* constrains one of the two parameters, ε or ω, and the other can be fitted; we choose to fit ε and therefore constrain ω. The second fixed parameter *μ*, the normal death rate, was calculated by taking the reciprocal of the average life expectancy (in days) of the 17 countries sampled, weighted by population size. We calculated a value of 80.3 years, based on data from individual countries (44). The third parameter *μ*_S_ is the sum of the normal death rate and an additional death rate due to a more severe form of the infection. We assumed a death rate of 3.8% (43) and that death follows after initial symptoms between 3 and 4 weeks (43). Thus *μ*_S_ = *μ* + 0.038/(3.5 * 7), where we use the average of 3 and 4 weeks and we convert to days with the factor of 7. The fourth fixed parameter *v*, the recovery rate once in the symptomatic state, was assumed to be the reciprocal of the average of 3 and 6 weeks (the range of recovery times) (4, 43) times the fraction of individuals in the symptomatic state that do not die, i.e. 1 - 0.038, so *v* = (1 - 0.038)/(4.5 * 7). The fifth fixed parameter *k*, the rate of viral decay in the environment, is the reciprocal of the average time that SARS-CoV-2 is expected to survive in the environment across a set of abiotic reservoirs, based empirical measurements (22). The sixth fixed parameter *p*, the fraction of individuals in the *I*_*A*_ state that move on to recovery without experiencing severe symptoms, was taken to be 0.956 (2).

We fit our model variations to the daily new cases data provided (See: supplementary information) starting on the day when there were ≥10 cumulative infected cases in that region. We choose the starting point of 10 cumulative cases in order to allow the outbreak to settle into a more consistent doubling time while also providing enough of an early-on window to capture the dynamics relevant to the ℛ_*0*_ and force of infection estimations.

We calculate the number of daily new infections in our model by numerically integrating the influx rate of new symptomatic infections over the course of a single day (i.e. ∫ *(1 - p) ω I*_*A*_ *dt*). We perform this calculation for each of 30 consecutive days and fit these values to the daily new cases data. We use the influx rate of symptomatic infections, as opposed to the total rate of new infection (including asymptomatic individuals), as we expect that the large majority of reported cases in the early COVID-19 outbreak to be symptomatic. And we expect that—in the outbreaks—almost all asymptomatic cases go unreported (20).

### Initial conditions

For each country, we use the first cumulative count that is ≥10 as a proxy for the initial number of *active* symptomatic cases *I*_*S0*_. We can justify this by proposing that, given that the doubling time is expected to fall between 3 and 6 days (45) then the exponential growth rate parameter of the infection (*r in exp(rt)*) would fall between 0.231 days^-1^ and 0.116 days^-1^ respectively. And, assuming 1 initial infected individual, the time to reach 10 cases for the former rate would be about 10.0 days (∼log(10)/0.231) and the time for the latter would be 19.8 days (∼log(10)/0.116). Thus, since recovery of symptomatic individuals, which takes between 3 and 6 weeks (39, 43) exceeds this interval, we expect that at the point when 10 cases have accumulated, all cases are still active. Lastly, in fitting the data to our model, we initialize all fitting parameters to a value of 1.5, in whatever units are appropriate for that parameter (expected to be close to the true value for most of the fitting parameters).

As an estimate for the initial number active *asymptomatic* cases, we take *I*_*A0*_ *= I*_*S0*_. That is, we expect that there are approximately as many asymptomatic cases as symptomatic cases early on. This assumption appears to be consistent with empirical findings. For example, data from the Diamond Princess cruise liner (11, 46), where all passengers were tested, revealed that approximately half of positively-testing cases were asymptomatic. Lastly, we assumed that the initial number of exposed individuals was approximately ℛ_*0*_*· (I*_*A0*_ *+ I*_*S0*_), based on the supposition that each of the initially infectious individuals *(I*_*A0*_ *+ I*_*S0*_) will have exposed the infection to approximately ℛ_*0*_ other individuals. We take the value of ℛ_*0*_ in this case to be 2.5, based on prior studies (18, 20). The *R* population is assumed to be 0 in the early stage of the outbreak, given the (average) 3 to 6-week recovery delay of COVID-19. The *W* population is assumed to be 1%.

### Basic reproductive ratios (ℛ_0_)

We can express the ℛ_*0*_ (eq. 7) in a form that makes explicit the contributions from the environment and from person-to-person interactions. In this way, the full ℛ_*0*_ is observed to comprise two ℛ_*0*_ sub-components: one the number of secondary infections caused by a single infected person through person-to-person contact alone (*R*_*p*_) and the other is the number of secondary infections caused by exchanging infection with the environment (*R*_*e*_).

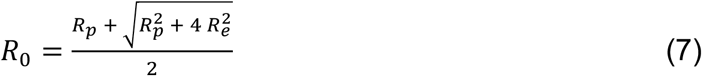

where *R*_*p*_ and *R*_*e*_ are defined in equations 8a and 8b.

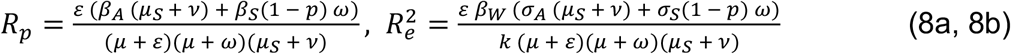

Note that when *R*_*p*_ = 0, the ℛ_*0*_ reduces to *R*_*e*_ and when *R*_*e*_ *=* 0, the ℛ_*0*_ reduces to *R*_*p*_. Thus, when person-to-person transmission is set to zero, the ℛ_*0*_ consists only of terms associated with transmission from the environment, and when transmission from the environment is set to zero, the ℛ_*0*_ consists only of infection directly between people. When both routes of transmission are turned on, the two ℛ_*0*_*-*components combine in the manner in equation 7.

While *R*_*e*_ represents the component of the ℛ_*0*_ formula associated with infection from the environment, the square of this quantity *R*_*e*_^*2*^ represents the expected number of *people* who become infected in the two-step infection process: people → environment → people, representing the flow of infection from people to the environment, and then from the environment to people. Thus, while *R*_*p*_ gives the expected number of people infected by a single infected person when the environmental transmission is turned off, *R*_*e*_^*2*^ gives the expected number of *people* infected by a single infected person by way of the environmental route exclusively no direct person-to-person transmission). Also note that *R*_*e*_^*2*^ */*(*R*_*e*_^*2*^ *+ R*_*p*_) can be used to measure the extent of transmission that is mediated by the environment exclusively. This proportion can be used as a proxy for how important environmental transmission is in a given setting. Elaboration on formulas 8a-b—and associated derivation-discussions—appear in the supplementary information.

## Results

The results section covers the following sets of tests and analyses:

1. The process through which parameters were estimated through fits to country-level outbreak data.
2. Sensitivity analysis, to discuss how variation in parameters influences key aspects of virus transmission dynamics.
3. An examination of how mathematical model incorporates environmental transmission. We discuss a calculation of the proportion of the transmission in a given setting can be attributed to environmental transmission.
4. Simulations of “reservoir world” scenarios, where the environmental transmission value is set to one of the environmental settings for which there are published findings (12). This is designed to identify how hypothetical settings comprising SARS-CoV-2 environmental transmission of a certain kind influences disease dynamics.

### Establishing features of environmental transmission using country outbreak data

Using the Akaike information criterion (AIC), SEIR models with an environmental compartment (SEIR-W) provide a strong relative fit to country incidence data. As discussed in the **Methods**, we compared the performance of models with (SEIR-W) and without (SEIR) environmental transmission across multiple countries to assess the role of environmental transmission in different contexts. Using the fitted parameters provided in Tables 1-3, and S1-S3, we calculate AIC values for the two mechanistic models: the standard SEIR model and the SEIR-W model. Table S4 displays the summary of the AIC values for each model-type fit to the first 30 days after the first day with total counts ≥10. In 10/17 countries (including 9/11 European countries), the SEIR-W model provided a better fit to the country data. In Fig. 1, we display the comparative individual country fit results for 4 of the countries with the fastest 30-day case growth rates— Spain, Italy, Iran, and Switzerland. The SEIR-W variant provides a better fit (significantly lower AIC score) than the standard SEIR model for all of these. Note that, as features of independent country epidemics are myriad and difficult to disentangle, several aspects independent of the model structure could explain the superior fit of the SEIR-W models. Results for additional country fits can be found in the supplementary information, Fig.S2.

**Fig 1.**
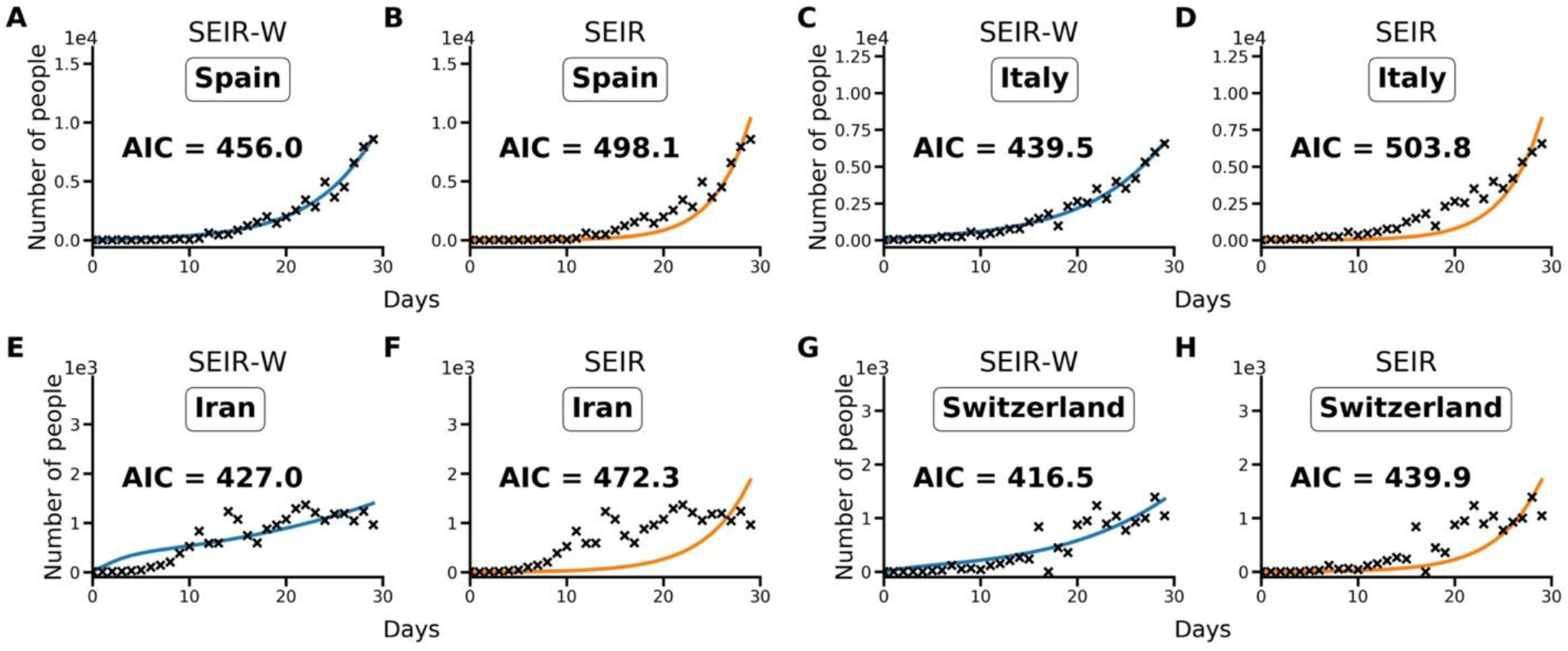
Illustrative model fit comparisons for SEIR-W and standard SEIR to case counts in early windows of the outbreak. The model fits are comparable across four countries with the largest early epidemics. These were chosen based having the highest cumulative number of infected cases after 30 days, following the first day when case counts were greater than or equal to 10. The four countries are (a,b) Spain, (c,d) Italy, (e,f) Iran and (g,h) Switzerland. These constitute a subset of 17 countries that had the highest number of cumulative COVID-19 cases (of the 181 total countries affected) as of March 30, 2020. Data come from the European Centre for Disease Control and Prevention, and from ourworldindata.org (40, 47). See supplementary information for more details.

### Sensitivity analysis reveals how environmental transmission can modulate disease dynamics

Partial Rank Correlation Coefficient (PRCC) analyses for the four examined features of the outbreak—(i) ℛ_0_, (ii) total number of infected individuals after 30 days, (iii) time to peak number of infected individuals, and (iii) size of peak number of infected individuals. Fig. 2 demonstrates the PRCC calculations for all four of these outbreak characteristics. For ℛ_0_, we observe that the model was strongly sensitive to several aspects related to virus transmission— *β*_*A*_, *β*_*S*_, *β*_*w*_——as well as the rate at which asymptomatic individuals develop symptoms (ω), the rate of recovery (*v*) and SARS-CoV-2 free-living survival rate (*k*).

**Fig 2.**
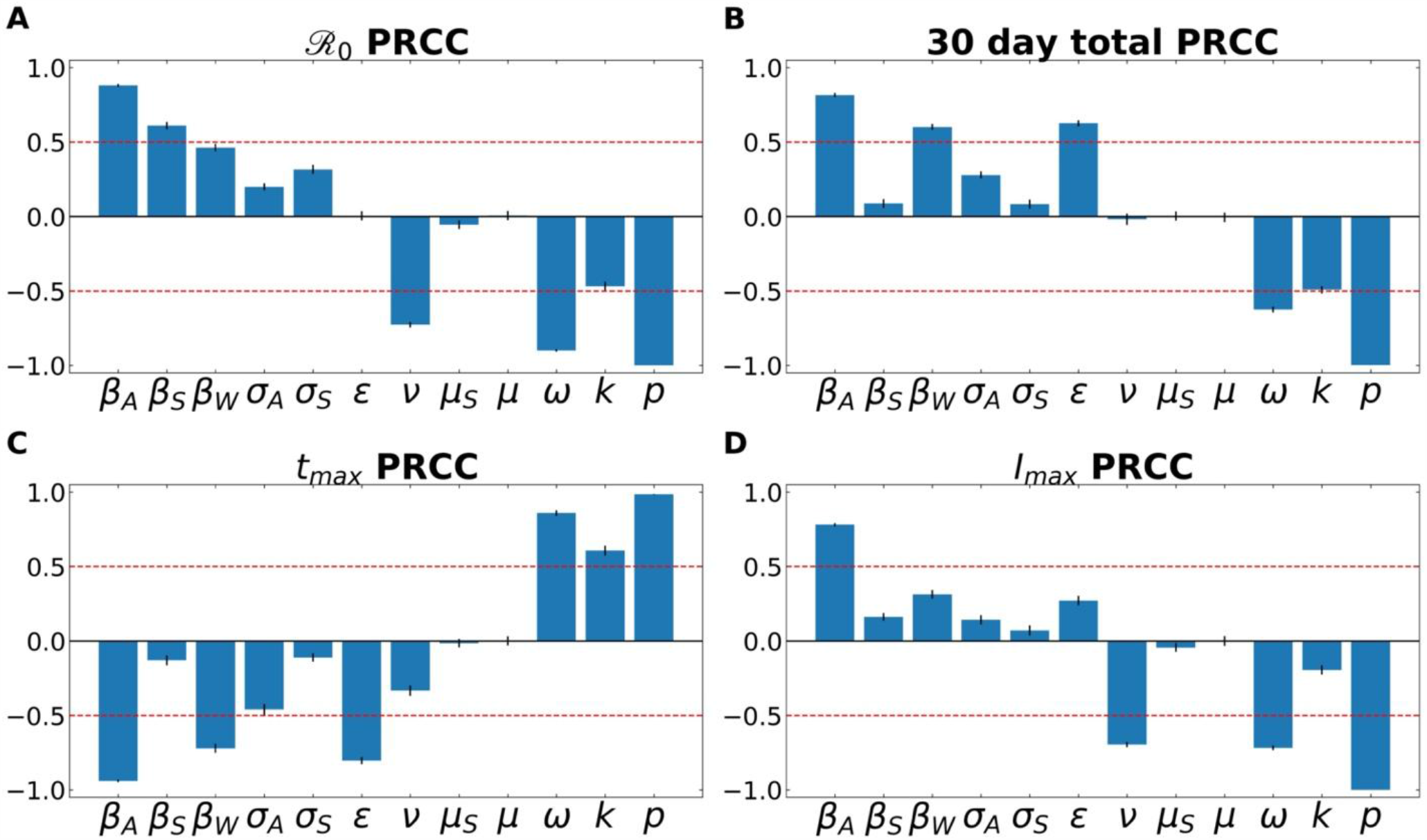
A Partial Rank Correlation Coefficient (PRCC) sensitivity analysis. PRCC was performed with respect to (A) ℛ_*0*_, (B) total number of infected (and symptomatic) after 30 days of outbreak, (C) time to peak number of symptomatic individuals (*t*_*max*_), and (D) peak number of symptomatic individuals. This analysis highlights the intercorrelated sensitivities of each of the model parameters. The blue bars show the mean value of each PRCC, with error bars at one standard deviation. This analysis was performed by sampling over uniform distributions of 4.5% around the nominal model parameter values. Parameters correspond to the fixed ones in Table 3, and the average fitted parameters values in Table S5. The red line marks PRCC values of +/- 0.50 and helps identify parameters that are more influential (greater than 0.50 or less than - 0.50). See supplementary information for more details.

One can also observe how some parameters are better suited to modify the peak of the infection, such as the recovery rate (*v*; which includes in it the swiftness of diagnosing and treating the virus). Others modulate the timing of the peak, such as *ε*, the rate of leaving the “exposed” compartment (or equally well, the reciprocal of the average time spent in the exposed compartment). Note that across all features, the fraction of cases that move along the “mild” route (*p*), from *E*→*I*_*A*_→*R*, has a powerful influence on all factors.

### The model emerging virus’ ℛ_*0*_ comprises person-to-person and environmental transmission

In the **Methods**, we described how the ℛ_*0*_ is composed of two sub-ℛ_*0*_ components, corresponding to different infectious interactions: person to person (*R*_*p*_), person to environment and environment to person (*R*_*e*_). Tornado plots were constructed that demonstrate how the ℛ_*0*_-components have their own architecture and sensitivity (Fig. 3).

**Fig 3.**
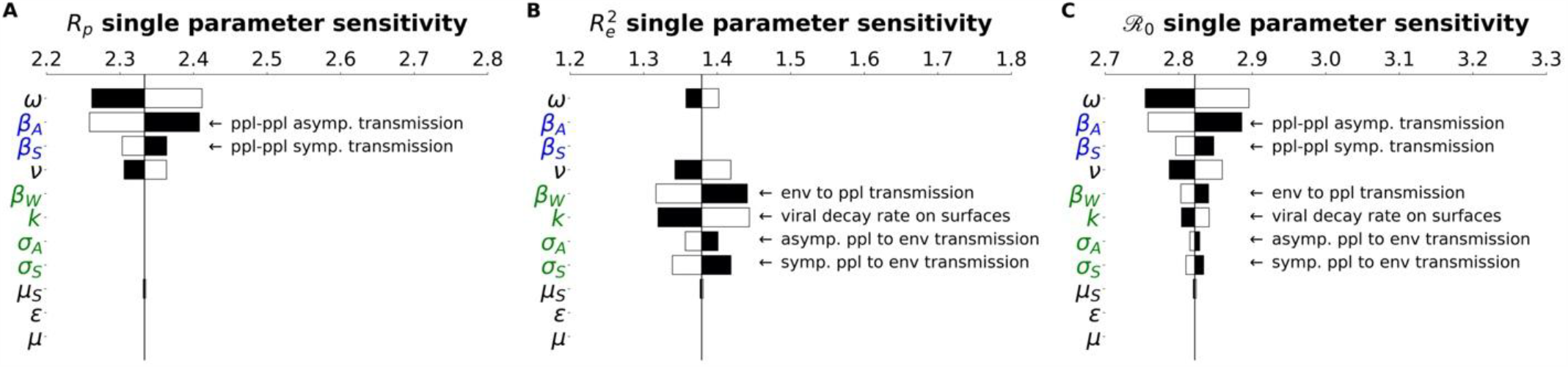
ℛ_0_ subcomponents have different parameter architecture. We compare the parameter architecture for the two ℛ_*0*_ components that compose the full ℛ_*0*_ expression, (a) *R*_*p*_, (b) *R*_*e*_^*2*^ and (c) ℛ_*0*._ Parameters are colored according to their relation with the environment or people: green parameters refer to the environment, blue parameters strictly refer to people, and black parameters are neutral in this regard. Black bars show the extent to which the component after changed when the parameter values are *increased* by 4.5%, The white bars show the same except for a *decrease* of 4.5%. For clarity, the single parameter that most influences the ℛ_*0*_ and its subcomponents is the faction of cases that move through the mild route (*p*) has been removed. For more details on how this parameter influences the ℛ_*0*_ and other features of the outbreak, see the PRCC analysis as discussed in the Methods and supplementary information.

In Fig. 4, we observe how variation in free-living survival (1/*k*) influences four characteristics of an outbreak: ℛ_*0*_, total number of infected individuals after 30 days, time to peak number of infected and symptomatic individuals, and maximum number of symptomatic individuals in the first 30 days. Note the annotations on the figure that highlight where the empirically-determined survival times of SARS-CoV-2 on a range of reservoir types (aerosol, copper, plastic, cardboard, stainless steel) (12). Also note that the quantitative relationships between 1/*k* and various outbreak features are slightly different. For example, the ℛ_*0*_ increases more gradually across a wider range of free-living survival values than some of the other features (Fig. 4).

**Fig. 4.**
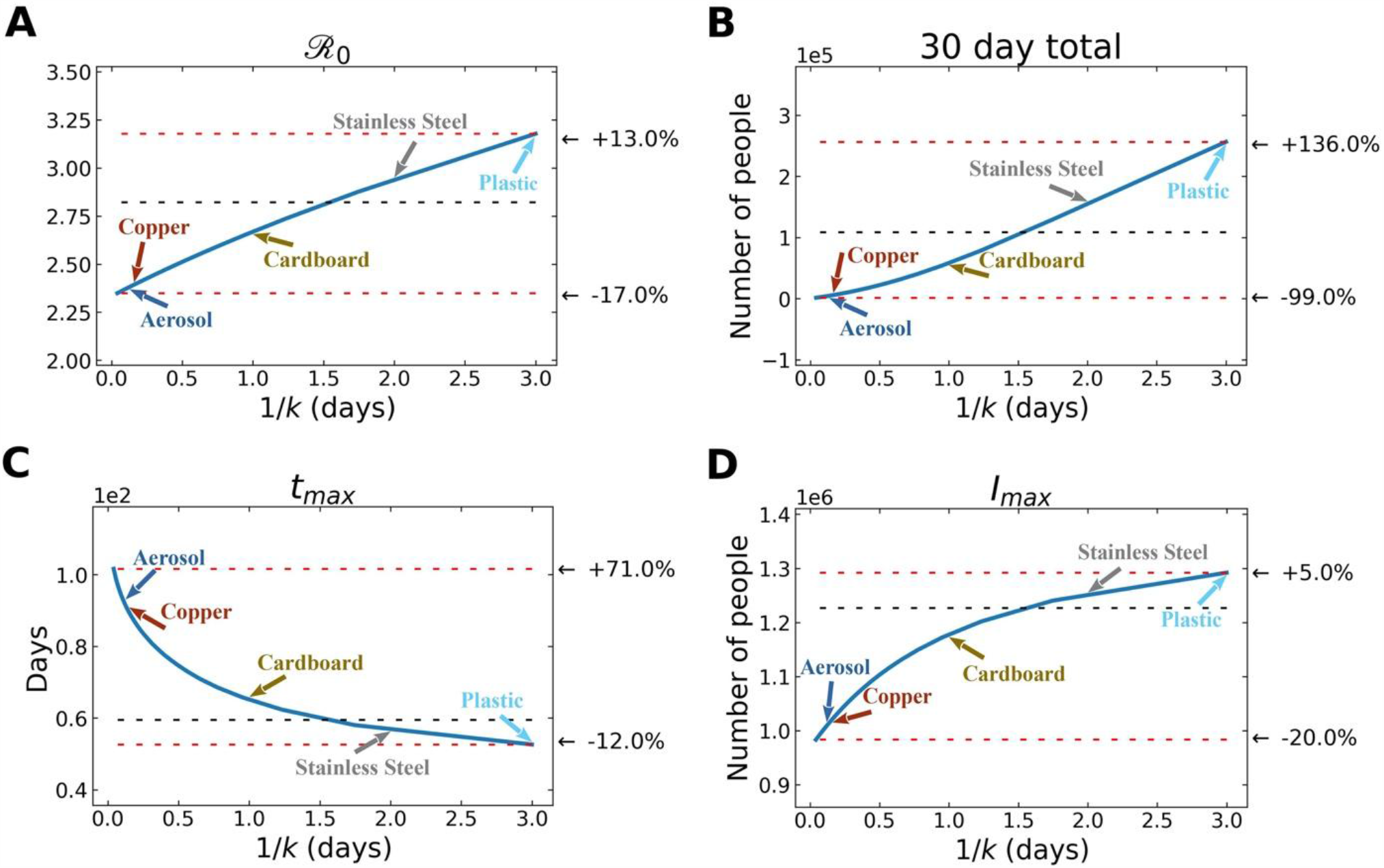
Environmental transmission: Various features of an outbreak change as a function of 1/*k*. (where *k* is the rate of decay of SARS-CoV-2 survival in the environmental compartment): to (A) ℛ_*0*_, (B) total number of infected (and symptomatic) after 30 days of outbreak, (C) time to peak number of symptomatic individuals (*t*_*max*_), and (D) peak number of symptomatic individuals. The black dashed lines show the value of the respective plotted value at the average value of 1/*k* (∼ 1.5 days), used in the fits from above. The top red line shows the maximum of plotted value for either the smallest value of 1/*k* chosen (*= 1 hr)* or the largest value of 1/*k* chosen (*= 3 days)*, depending on whether the plotted value decreases or increases with 1/*k*, and the bottom red line shows the plotted value at the other extreme of 1/*k*. We emphasize that these correspond to indirect, environmental transmission only. Aerosol transmission, for example, is likely a cause of direct transmission between individuals.

We should reemphasize some aspects of the underlying physics of the simulations in this study that were introduced in the Methods section. In reality aerosol transmission likely contributes to person-to-person transmission. There, however, scenarios where aerosols serve as environmental reservoir, capable of transmitting between individuals in an “indirect” way. In this study, we use it in an analogous way to surfaces, where air may be exchanged in the same room where infected individuals were, rather than exchanging infectious particles on a surface.

### The composition of abiotic reservoirs modulates outbreak dynamics

Fig. 5 depicts the results of “reservoir world” simulations, where the *k* values correspond to those from a 2020 study highlighting the survival of SARS-CoV-1 and SARS-CoV-2 on different physical surfaces (12). The summary of these simulations (Fig. 5) highlights that the surface composition of a setting has a meaningful impact on several features of outbreak dynamics. Of the “reservoir world” simulations, “aerosol world” (5a) and “copper world” (Fig. 5c) takes the longest amount of time (91.5 and 88.4 days, respectively) to rise to the peak number of infected-symptomatic individuals, indicating an outbreak which is slower to develop. Relatedly, the ℛ_*0*_ values are much different in the different “reservoir world” scenarios: The “aerosol world” simulation has an ℛ_*0*_ of 2.38, the “copper world” simulation an ℛ_*0*_ of 2.4, and the “plastic world” simulation an ℛ_*0*_ of 3.18 (Fig. 6 and Table S5). In addition, the total number of individuals infected after 30 days of the outbreak, and the total number dead after 30 days are both significantly lower in the “aerosol world” and “copper world” setting (Fig. 6 and Table S5). The peak value of infected individuals is not dramatically different across “reservoir worlds.” That is, while many features associated with severity differ greatly across “reservoir world” settings, we observed significantly less variation in the peak of the epidemic as compared with the time to the peak of the epidemic (Table S5). Maybe the most noteworthy of the differences is the vast disparity in the number of deaths in the first 30 days of the outbreak, where the “plastic world” setting has more than 30 times the number of deaths as the “copper world” scenario (1,814 vs. 55, respectively; Fig. 6 and Table S5).

**Fig. 5.**
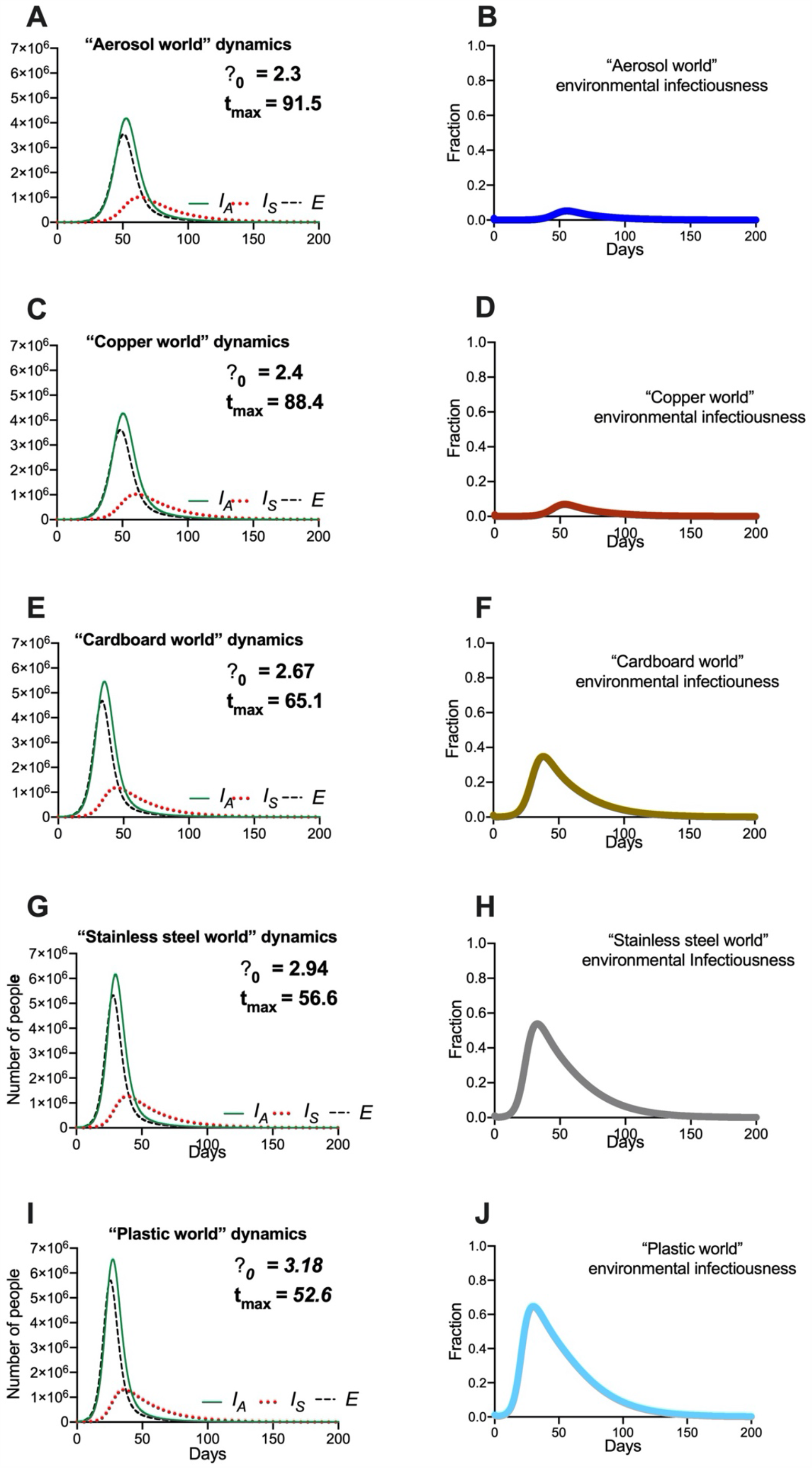
Hypothetical “reservoir world” simulations feature differing environmental transmission dynamics. Population and environmental dynamics of SEIR-W model outbreaks in hypothetical settings composed of pure substances where SARS-CoV-2 can survive and be transmitted. (A, B) “aerosol world,” (C, D) “copper world,” “(E, F) cardboard world,” (G, H) “stainless steel world,” and (I, J) “plastic world.” Environment infectiousness corresponds to the proportion of the environment that contains infectious SARS-CoV-2. Note that the surface where the viral decay is strongest (Copper), the peak of the epidemic is pushed farthest from the origin. Also note the ℛ_*0*_ values graphs A, C, E, and G, which highlight that the different “reservoir worlds” behave like fundamentally different outbreaks in several ways.

**Fig. 6.**
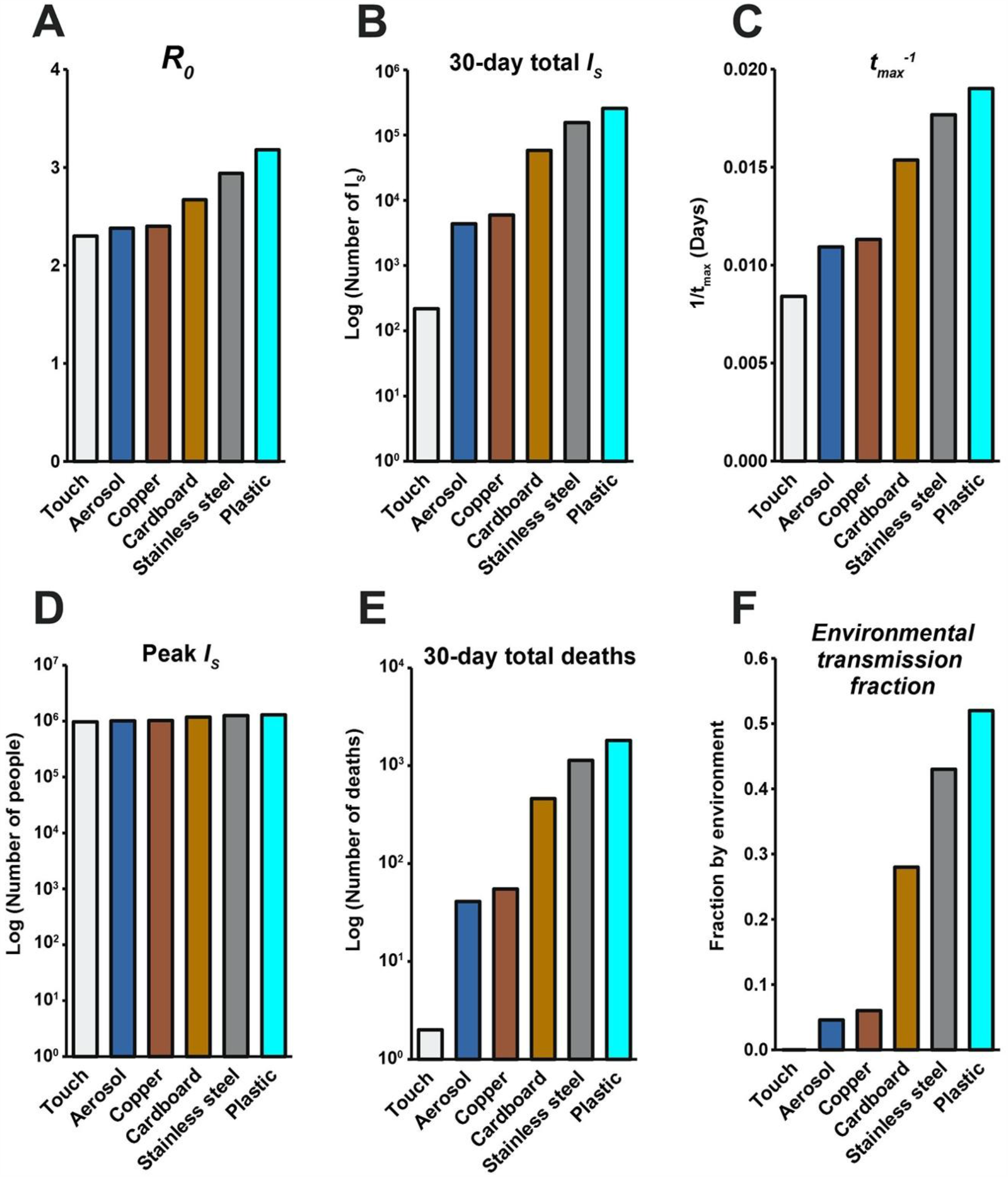
Summary of the hypothetical “reservoir world” outbreak intensity measures. *Graphs* correspond to the attributes of simulated epidemics where environments are entirely composed of a given physical surface, and larger values correspond to various aspects of outbreak intensity. A) ℛ_*0*_, (B) total number of infected (and symptomatic) after 30 days of outbreak, (C) the inverse time to peak number of symptomatic individuals (*t*_*max*_^*-1*^; larger values = shorter times to reach peak), (D) peak number of symptomatic individuals, (E) deaths after 30 days, and (F) environmental transmission fraction. Note the log scales on the y-axis in (B), (D) and (E).

Lastly, comparisons of the metric, *R*_*e*_^*2*^ */*(*R*_*e*_^*2*^ *+ R*_*p*_), which measures the extent to which transmissions can be attributed to the environmental route, further highlights how different environmental reservoirs influence disease transmission (Fig. 6). The “aerosol world” and “copper world” settings comprise 4.6% and 6% of transmission events occurring through non-person to person transmission. This differs dramatically from the “plastic world” setting, where 52% of transmission events are occurring through the environmental route.

## Discussion

### Deconstructing the basic reproductive number (ℛ_*0*_) into subcomponent reveals the role of environmental transmission

By deconstructing the basic reproductive number into components, we can better understand how variation in the ℛ_*0*_—by setting, time, or geography—may reside in how these contexts are driven by environmental transmission. Many of these effects may be (as they are in this study) localized to one component of the ℛ_*0*_, labeled *R*_*e*_^*2*^ in this study. Notably, the *R*_*e*_^*2*^ component is highly sensitive to the transmission interaction between people and the environment (β_w_), and the decay rate of virus in the environment (*k*). Interestingly, the *R*_*e*_^*2*^ is relatively robust to the rate of infectious virus shed into the environment from the asymptomatic infected individuals (the parameter called *σ*_*A*_ in this model). Also, deconstructing the ℛ_*0*_ value into these components facilitates the creation of new metrics that quantify how much a given epidemic is driven by certain routes. As many viral diseases may contain multiple transmission routes, being able to properly quantify their relative contribution may be useful for public health interventions.

### Determining the role of viral free-living survival on disease transmission should be the focus of early inquiry in the ecology of any emerging infectious disease

In this study, we introduce a general framework for studying the joint effects of direct and indirect microparasite transmission and then analyzed that model using data from SARS-CoV-2. We demonstrate that understanding the particulars of environmental transmission, including variation in viral free-living survival, can alter fundamental characteristics of disease dynamics. For SARS-CoV-2, we find that across 17 countries, the SEIR-W model better explained the epidemiological patterns than did SEIR models. Critically, the SEIR model included asymptomatic/sub-clinical transmission. Given that features of environmental transmission can influence central properties of disease dynamics, we should consider the possibility that variation in free-living survival may contribute to variation in aspects of disease dynamics. Nevertheless, that the SEIR-W model fits certain country data relative to others may be the consequence of many characteristics of an epidemic (e.g. quality of data, testing capacity), none of which represents anything meaningful about the mechanism of an outbreak. Indeed, the model is strongly sensitive to several aspects related to virus transmission, including the rate at which both symptomatic and asymptomatic individuals transmit infection to susceptible hosts, the rate at which asymptomatic individuals develop symptoms, the rate of recovery (*v*) and SARS-CoV-2 decay rate (*k*).

### Disease dynamics in different hypothetical “reservoir world” settings resemble essentially different outbreaks

Analysis of the ℛ_*0*_ and its subcomponents highlights that many aspects of outbreak dynamics are sensitive to the parameter associated with environmental decay rate (*k* in the model presented in this study). Analysis of hypothetical settings purely comprising reservoirs of a certain kind (“reservoir world”) fortifies the significance of free-living survival on physical surfaces and environmental transmission in outbreak dynamics. While our findings cannot speak to the outbreak dynamics in any particular setting in the real world, they do reveal that the surface composition of a setting can significantly influence the behavior of an outbreak. For example, the ℛ_*0*_ in the “plastic world” simulation (ℛ_*0*_ *=* 3.18) is over 1.3 times the ℛ_*0*_ in the “copper world” simulation (ℛ_*0*_ *=* 2.4). Many other differences between these outbreaks come as a consequence of the different ℛ_*0*_ values. For example, the “plastic world” simulation reaches a peak number of symptomatic infectious individuals almost 1.7 times faster than the “copper world” simulation, and kills over 30 times more people in the first 30 days (1,814 deaths in “plastic world” vs. 55 deaths in the “plastic world”).

These differences are so significant that they might be naively interpreted as completely different outbreaks early on in an outbreak. Note, however, that the maximum value of the infected-symptomatic populations are roughly equivalent across “reservoir worlds,” and so the influence of SARS-CoV-2 survival on physical surfaces (mediated by difference in free-living survival) doesn’t affect all aspects of outbreak dynamics equally.

Despite the breadth of differences observed across surfaces, a very notable finding regards the similarity between the “aerosol world” and “copper world” results. That is, an outbreak in a hypothetical world where there are no physical surfaces, but only transmission via aerosols would be only slightly more intense than an outbreak where indirect transmission was driven entirely by copper (using ℛ_*0*_ as a quick proxy, “aerosol world” = 2.38, and “copper world” = 2.4). The implications here are subtle, but worth elaborating on: while a lot of public debate has focused on a dichotomy between aerosol-mediated transmission and environmental transmission, our findings suggest that the differences between aerosol transmission and some physical surfaces is so minute that the epidemiological signature for differences between them may be indistinguishable. Consequently, the more productive debates would focus, not on whether environmental transmission is occurring at all, but how the combination of aerosols and surfaces contribute to non-contact transmission events.

### Implications for the ecology of emerging outbreaks

As of June 1, 2020, the scientific community remains in the fact-finding phase of SARS-CoV-2 biology and COVID-19 understanding. A significant source of fear and speculation in the pandemic involves the plausibility that SARS-CoV-2 has undergone local adaptation in certain settings, translating to different epidemiological properties. While there is no currently convincing molecular or clinical support for local adaptation in SARS-CoV-2, our findings highlight how easy it is to conflate an environmental (or ecological) difference for a genetic one: the same virus, spreading in populations of identical size and behavior, differing only in the composition of physical surfaces where the virus can be transmitted through the environment, can have ℛ_*0*_ values between 2.4 and 3.18, with early death rates 30 times apart. Even further, our findings highlight how models of direct and indirect/environmental transmission can provide comparable results, rendering it easy to conflate routes of transmission. This is related to a problem known as the identifiability problem that arises when models are fit to early outbreak data (48).

These difficulty in predicting outbreaks conferred by this identifiability problem, and the generic variability in disease dynamics from setting to setting has been encapsulated in a concept called permutation entropy (49). Even more, environmental reservoirs composed of certain physical surfaces (plastic-like in our model) may be associated with phenomenon resembling a “superspreading” event, where individual variation in contagiousness can drive unusually large numbers of infections (50). Perhaps a better understanding of how, and on what surfaces, viral populations survive may one day improve the predictability of outbreak trajectories.

Because of the identifiability problem, and a general lack of consensus regarding the mechanisms of transmission in various diseases, we caution against the overextrapolation of these findings to any particular epidemic. As of July 2020, the evidence for widespread surface transmission of SARS-CoV-2 remains dubious. Insofar as SARS-CoV-2 is not the last of the emerging infectious diseases, then we should remain vigilant about understanding how different routes of transmission may influence disease dynamics. This is especially true because there remain other microparasites (viral and other) in human and nonhuman hosts that are transmitted via the environmental route that will continue to require our attention.

## Data Availability

Data are either available or the source is referenced in the main text and supplemental information. The code used for the analyses in this study is publicly available on Github: https://github.com/OgPlexus/Copperland

## Acknowledgments

The authors would like to thank W. Turner, L. Smolin, S. Ramachandran, and D. Weinreich for seminar invitations where iterations of the ideas in this manuscript were discussed. The authors would also like to thank J. Weitz for feedback on an earlier draft of the manuscript. ALM would like to acknowledge NIH U54GM115677 for funding support. The authors report no conflicts of interest.

## Data availability

All data is available in the main text or the supplementary information. The code used for our analyses is publicly available on Github: https://github.com/OgPlexus/Copperland

## Notes

### Competing Interest Statement

The authors have declared no competing interest.

